# The Relation Between Viscous Energy Dissipation And Pulsation For Aortic Hemodynamics Driven By A Left Ventricular Assist Device

**DOI:** 10.1101/2022.07.12.22277566

**Authors:** Akshita Sahni, Erin E. McIntyre, Kelly Cao, Jay D. Pal, Debanjan Mukherjee

## Abstract

Left ventricular assist device (LVAD) provides mechanical circulatory support for patients with advanced heart failure. Treatment using LVAD is commonly associated with complications such as stroke and gastro-intestinal bleeding. These complications are intimately related to the state of hemodynamics in the aorta, driven by a jet flow from the LVAD outflow graft that impinges into the aorta wall. Here we conduct a systematic analyses of hemodynamics driven by an LVAD with a specific focus on viscous energy transport and dissipation. We conduct a complementary set of analysis using idealized cylindrical tubes with diameter equivalent to common carotid artery and aorta, and a patient-specific model of 27 different LVAD configurations. Results from our analysis demonstrate how energy dissipation is governed by key parameters such as frequency and pulsation, wall elasticity, and LVAD outflow graft surgical anastomosis. We find that frequency, pulsation, and surgical angles have a dominant effect, while wall elasticity has a weaker effect, in determining the state of energy dissipation. For the patient-specific scenario, we also find that energy dissipation is higher in the aortic arch and lower in the abdominal aorta, when compared to the baseline flow without an LVAD. This further illustrates the key hemodynamic role played by the LVAD outflow jet impingement, and subsequent aortic hemodynamics during LVAD operation.

## 1 Introduction

Mechanical circulation support (MCS) in the form of Left Ventricular Assist Devices (LVADs) has emerged as a primary treatment modality for advanced heart failure patients, both as a destination therapy and bridge-to-transplant^[38]^. Within the United States alone, more than 6 million individuals over the age of 20 have advanced heart failure^[4]^. Despite advancements in LVAD design and therapy, patient outcomes on LVAD support remain associated with significant morbidity and mortality due to severe complications such as stroke and GI bleeding^[28,1,27,35]^. It is widely acknowledged that the altered state of hemodynamics post-LVAD implantation, as compared to the baseline physiological flow pre-implantation, has an intimate connection to treatment efficacy and underlying complications. Advancements in understanding spatiotemporally varying hemodynamic features can be critical for treatment efficacy assessment^[3,37]^. This has motivated a wide range of studies on characterization of hemodynamics post-LVAD implantation, including investigations on: (a) LVAD outflow graft surgical attachment angle and its influence on thromboembolic risks ^[2,30]^; (b) influence of additional surgical parameters^[5]^; (c) role of pulse modulation^[6]^; and (d) effect of intermittent aortic valve reopening^[25]^. Existing studies have mainly looked into factors like flow stasis, recirculation, mixing, and state of wall shear. One aspect that has remained sparsely investigated in the context of LVAD therapy is flow energetics and energy dissipation, commonly quantified in terms of a viscous dissipation rate (VDR). Viscous dissipation has been previously used to quantify flow efficiency and correlate with patient outcomes in reconstructive surgical procedures such as Fontan procedure^[7,44]^ and variants with Total Cavopulmonary Connection(TCPC)^[15,24,9]^. Energy dissipation has also been investigated as a descriptor for ventricular hemodynamics and cardiac function^[11,12]^. In terms of Ventricular Assist Devices design, energy dissipation has been discussed through the role of high shear and turbulence in blood damage as blood moves through the pump, looking at energy dissipation as a turbulence parameter^[13,18]^. However, details of viscous dissipation for hemodynamics during LVAD operation, and its interplay with ventricular-aortic coupling, remain incompletely understood. Here we present a systematic computational hemodynamics study for viscous energy dissipation, comprising: (a) parametric investigations of dissipation as function of frequency, pulsation, and wall elasticity for flow through idealized cylindrical tube models of single vessels; and (b) parametric investigations of dissipation as function of surgical parameters and pulsation for flow through a patient-specific vascular anatomy with attached LVAD outflow graft.

## 2 Viscous dissipation analysis in idealized cylindrical vessels

### 2.1 Viscous dissipation rate in incompressible Newtonian flow

Here, we briefly reproduce the mathematical expressions for quantifying viscous dissipation rate (VDR) for an incompressible Newtonian fluid flow. The generalized equation of energy balance is stated as follows:

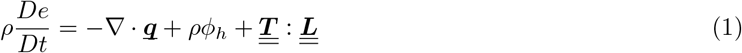

where *ρ* is the fluid density, *e* denotes internal energy density, <u>***q***</u> denotes thermal fluxes in the flow, *ϕ*_*h*_ denotes additional heat sources or sinks, 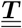 denotes the total fluid stress tensor, and 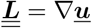 is the velocity gradient tensor. In the absence of thermal contributions and sources/sinks, energy losses are induced by mechanical deformation captured in the last term in Equation 1. The velocity gradient can be further decomposed as:

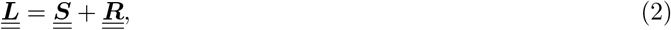

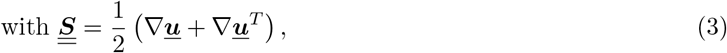

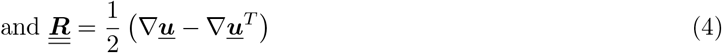

where 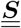 is the symmetric strain-rate tensor, and 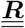 denotes the anti-symmetric spin tensor for the flow. Assuming that the fluid obeys angular momentum conservation and a Newtonian constitutive relation, we can utilise the symmetry of the fluid stress tensor 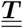 to subsequently obtain the following expression for the rate of deformation work per unit volume in the flow:

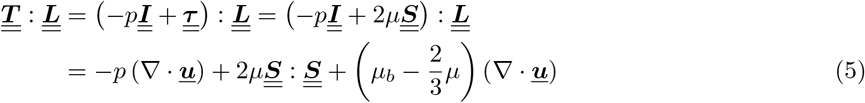

For the case of incompressible flows with zero divergence, Equation 5 above reduces to mechanical work due to viscous stresses alone, leading to the definition of the viscous dissipation rate *ϕ*_*v*_ as follows:

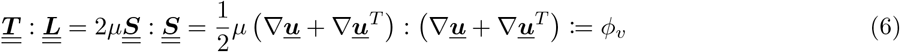

The energy balance equation can be stated in terms of viscous dissipation rate *ϕ*_*v*_ as follows:

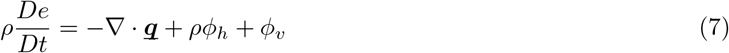

We note that for non-Newtonian incompressible flows, the appropriate constitutive relation can be incorporated in Equation 5 to derive (wherever appropriate) expressions for the corresponding viscous dissipation rate *ϕ*_*v*_. Here, we focus strictly on Newtonian fluids, leveraging the widely employed assumption that blood in the large arteries behave nearly as a Newtonian fluid^[22,39]^.

### 2.2 Viscous dissipation in rigid cylindrical vessels

Based on the fundamental definitions presented in Section 2.1, here we briefly revisit the theory for analyses of VDR for pulsatile flows through a rigid cylindrical tube of circular cross-section. We directly follow theoretical analysis outlined in classical works^[45,46,14]^, reproducing key expressions for our study. Specifically, we consider purely axisymmetric axial flow with velocity *u* in the axial *x*-direction, driven by a pulsatile pressure gradient *k* = *dp/dx* which can be decomposed into a mean or steady component, and an oscillatory component; leading subsequently to a mean and oscillatory flow velocity as shown below:

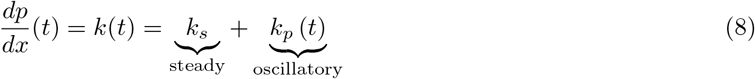

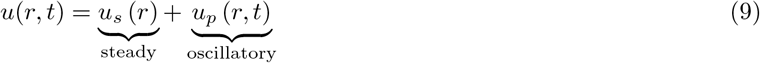

The mean flow component *u*_*s*_ will resemble classical Poiseuille flow in cylindrical tubes, with the axial flow velocity *u* obeying the following relation:

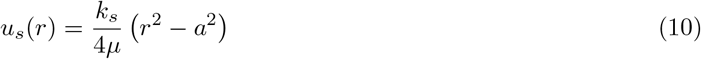

where *a* is the tube circular cross-sectional radius, and *μ* is dynamic viscosity of the fluid (blood, in this case). Likewise, assuming that the oscillatory pressure gradient component is of the form *k*_*p*_ = *k*_0_*e*^*iωt*^ (*ω* being oscillation frequency), the corresponding oscillatory flow velocity solutions can be assumed similarly as *u*_*p*_ = *U*_0_*e*^*iωt*^; which, when plugged in to the mass and momentum balance equations leads to the following solution for oscillatory axial velocity as derived in ^[45,46]^:

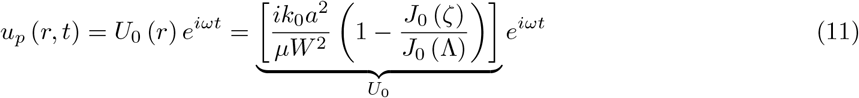

where we have:

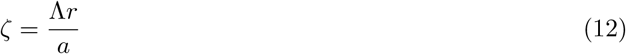

and the variable Λ is further related to the flow Womersley Number *W* as:

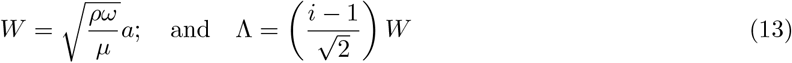

With these expressions, and using the definition of VDR as outlined in Section 2.1, we can obtain expressions for the VDR (*detailed derivation not shown, please refer to details in Supplementary Material*). The VDR contribution solely from the steady/mean contribution to the flow can be obtained as:

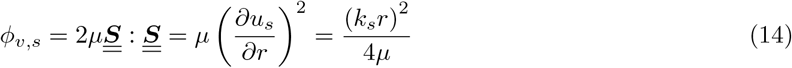

Similarly, the expression for the VDR obtained solely from the oscillatory part of the flow field, can be obtained as:

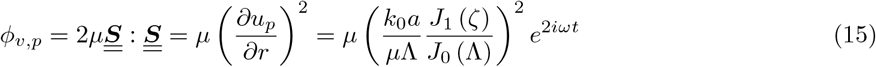

Given the total flow velocity is a combination of *u*_*s*_ and *u*_*p*_, the net VDR for the total flow can be obtained as:

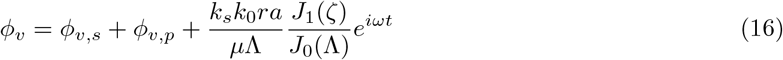

For generalized arbitrary pulsatile flow waveforms, the pressure gradient will be decomposed into a steady component and a series of oscillatory components of varying frequencies *ω* using Fourier transform, leading to an overall *ϕ*_*v*_ which will include the influence of each frequency component, in a form as stated below:

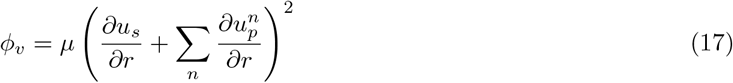

where *n* denotes summation over the different frequency contributions obtained from Fourier decomposition. For additional details please refer to the supplementary material.

### 2.3 Viscous dissipation in elastic cylindrical vessels

Here, we briefly revisit the theory for analyses of pulsatile flows through an elastic cylindrical vessel, reproducing key expressions from classical works^[45,46,14]^ for our study. For elastic vessels, matching boundary conditions at the moving wall of the tube leads to both axial velocity *u*(*x, r, t*) and radial velocity *v*(*x, r, t*) that vary along the axis and along cross-section of the vessel. Elastic deformation of vessel wall leads to propagation of a wave along the axis of the vessel. For a pulsatile pressure gradient *dp/dx* driving flow through a cylindrical tube, it is commonly assumed that: (a) the length of the propagating wave is much larger than tube mean radius *a*; and (b) the characteristic average flow velocity is much smaller than the characteristic wave propagation speed (Korteweg Moens wave speed). These assumptions lead to several simplifications for the governing equations of mass and momentum balance, and similar to the analysis in Section 2.2, we can decompose the corresponding pressure and velocity solutions into a mean (or steady) and oscillatory components. However, unlike rigid vessels, these oscillatory solutions must account for wave propagation along the axial (*x*) direction. This leads to the following form of oscillatory pressure and velocity components (with the understanding that the steady component will yield an equivalent Poiseuille flow solution as also outlined in Section 2.2):

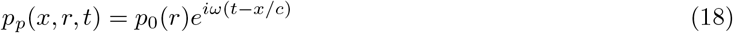

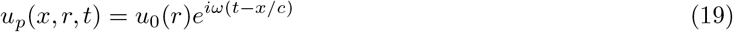

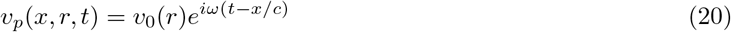

where *c* denotes the effective wave propagation speed in the vessel accounting for fluid viscosity (different from the inviscid Korteweg-Moen speed). We assume further that the oscillatory pressure gradient is constant across the vessel cross-section with a magnitude *B* such that:

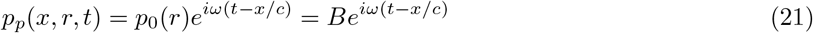

Using the solutions in Equations 18-21, substituting them in the simplified governing equations for mass and momentum balance, and assuming that deformation of vessel wall obeys linear elastic mechanics, the following forms of the oscillatory flow velocity components have been obtained in prior classical texts^[45,46]^ (details of derivation not reproduced here):

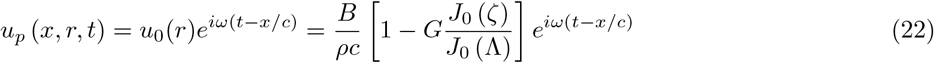

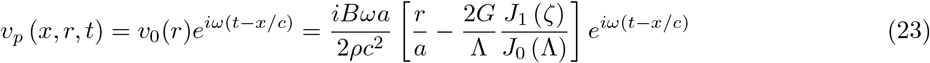

The effective viscous wave speed *c* is determined based on an algebraic equation in terms of an intermediate algebraic parameter *z* such that:

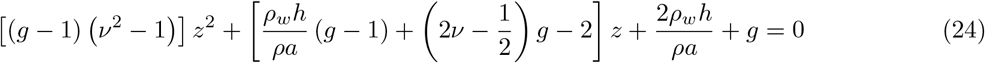

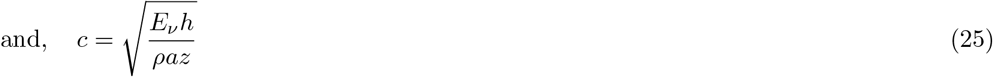

where, *ρ*_*w*_ is vessel wall density, *ν* is Poisson’s ratio, *ζ* is the same as defined in Equation 12; *g* is a frequency dependent parameter defined in terms of Womersley number *W* as follows:

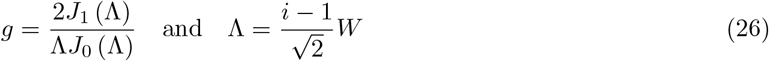

and, the term *E*_*ν*_ is defined as:

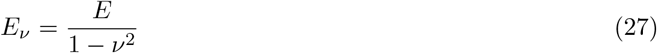

where *ν* is the Poisson ratio for the vessel wall material. Lastly, the parameter *G* in the expressions outlined above is commonly referred to as the elasticity factor, and defined as:

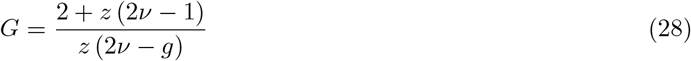

With these set of expressions for the state of flow in the tube, the VDR estimates can be computed for both the steady and the oscillatory components. The VDR from steady component alone (*ϕ*_*v,s*_) is the same as for the case of rigid tubes defined in Equation 14. The VDR from oscillatory component alone can be computed by plugging in the expressions for *u*_*p*_(*x, r, t*) and *v*_*p*_(*x, r, t*) outlined in Equations 22 and 23 in the expression for VDR, and combining both axial and radial contributions as follows:

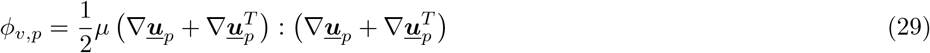

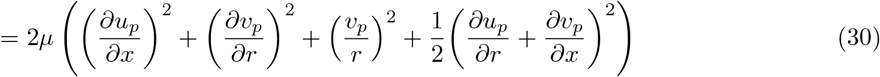

where further detailed algebraic expressions are not provided, but included in the Supplementary Material. Finally, similar to the case of rigid tubes, for a generalized arbitrary pulsatile pressure gradient driving the flow, we can decompose the waveform into a steady component and a seres of oscillatory components of varying frequencies using Fourier decomposition, and the overall *ϕ*_*v*_ can be computed based on contribution from each frequency as follows:

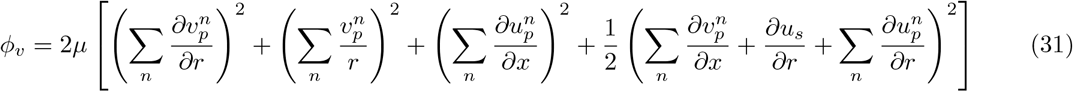

where *n* denotes summation over the different frequency contributions obtained from Fourier decomposition. For additional details please refer to the supplementary material.

### 2.4 Numerical experiments on idealized cylindrical vessels

We designed a numerical study for systematic parametric investigations on VDR in pulsatile flows, using idealized cylindrical tubes. A schematic overview of the numerical study design is shown in panel a. in Figure 1. We considered two different cylindrical tube cases. The first case comprised a tube of diameter 0.006 m, which is equivalent of the human common carotid artery. The second case comprised a tube of diameter 0.02 m, which is equivalent of the human aorta. These two representative segments were chosen because of the key role these two vascular regions play in LVAD related complications. For each case, four different wall elasticity values were chosen: E = 100 kPa; 1 MPa; 10 MPa; and ∞ (which corresponds mechanically to a rigid walled vessel). These elasticity values were assigned based on ranges of measured values for the carotid artery and the aorta reported in the literature^[17,47,21]^. Further, for each wall material choice, a set of controlled flow pulsations were considered.

**Figure 1:**
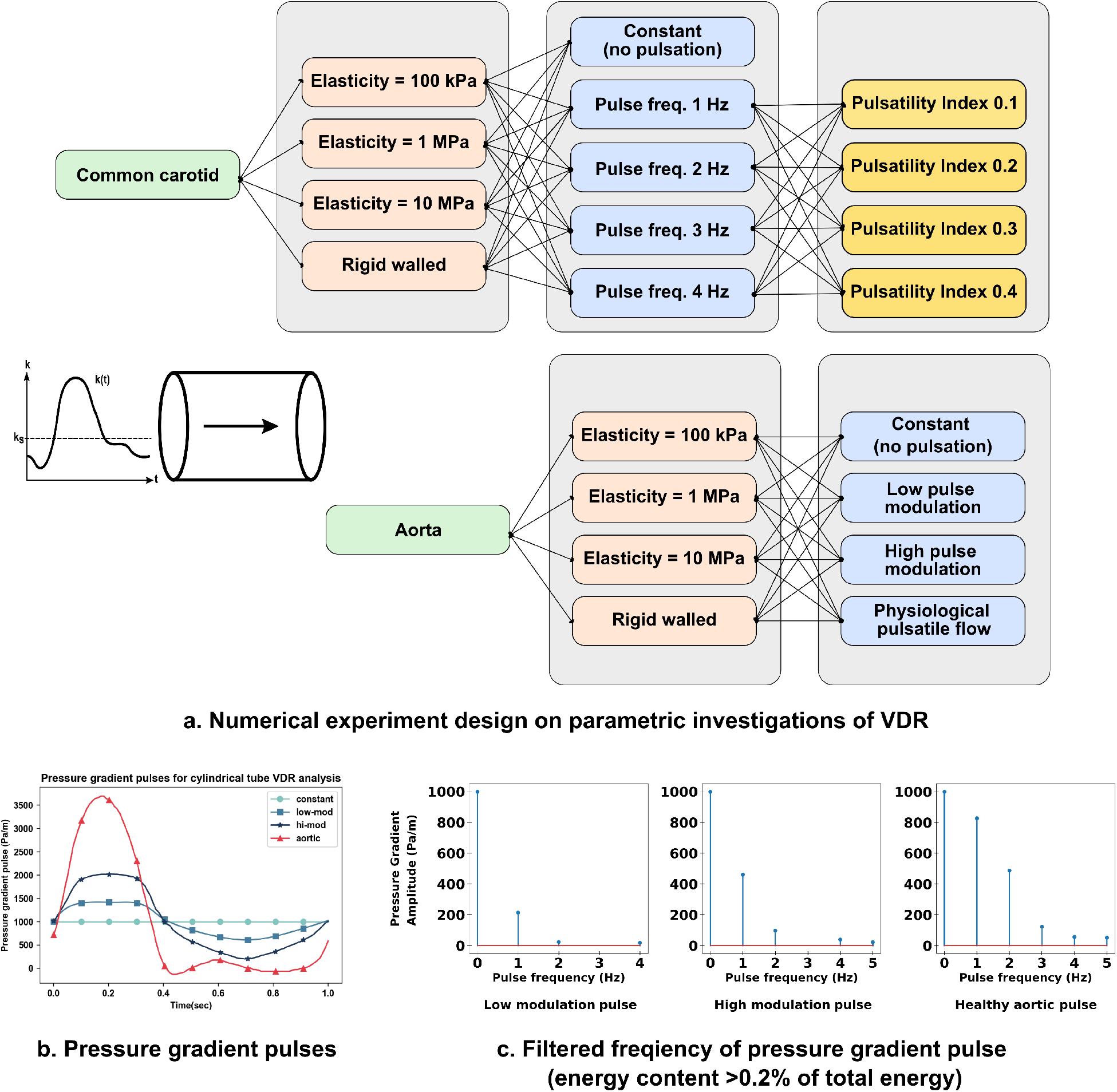
Illustration of study design for parametric investigations of viscous dissipation rates in idealised cylindrical vessels. Panel a. depicts the numerical experiment design details, panels b. and c. depict the pressure gradient pulses used for aortic equivalent vessel and their filtered frequency compositions respectively.

For the carotid equivalent tube, five different pulsatile pressure gradient profiles with an average (*k*_*av*_) of 550 *Pa/m* were imposed to drive the flow, using the following relation:

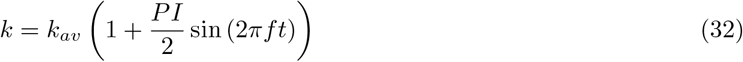

The average value of these pulses (*k*_*av*_) was matched with the average pressure gradient derived from physiologically observed common carotid flow rate ranges and artery resistance^[41]^. We chose: (a) a constant gradient (that is, zero frequency); and (b) sinusoidal pulse frequencies (*f*) 1 Hz, 2 Hz, 3 Hz and 4 Hz. The selected frequencies were identified based on a Fourier series decomposition of measured common carotid artery flow profiles reported in literature^[20]^. Pulsatility indices (PI) of 0.1, 0.2, 0.3 and 0.4 were prescribed for each sinusoidal pulse with non-zero frequency. This resulted in 17 different combination of pressure-gradient profiles driving flow through the tube. Corresponding Womersley numbers are W = 4.5, 6.4, 7.8 and 9.0 respectively for each of the sinusoidal pulse cases considered here.

For the aortic equivalent tube, four different pulse modulation scenarios were considered. Unlike the carotid tube model, these were chosen to be representative of pulsatile flow profiles typically observed in the aorta under healthy and disease scenarios - corresponding to ventricular dysfunction and mechanical circulatory support with low and high extent of pulse modulation derived from existing literature^[16]^. Each of these profiles were scaled to ensure the same mean flow of 5.0 L/min is driven through the vessel (refer Table 1 for parameter details). The four resulting pressure gradient profiles are illustrated in panel b. in Figure 1. We analyzed the filtered frequency contributions within all the pulses and observed that frequencies (*f*) 1 Hz, 2 Hz and 4 Hz are dominant driving frequencies as shown in panel c. Figure 1. This further supplements the rationale behind using the above single frequency pulses for the carotid equivalent tube analysis. For each pulse considered here, the Womersley numbers were calculated using maximum filtered frequencies as: W = 33.3 for the physiological aortic pulse profile and high pulse-modulated flow profile; and W = 29.8 for the low pulse-modulated flow profile.

**Table 1:** A table of input parameter values used in the numerical experiment design for pulsatile flow in common carotid and aorta sized idealised cylindrical vessels

| Cylindrical Tube | Length<br>(m) | Diameter<br>(m) | Thickness<br>(m) | Resistance<br>(Pa-s-m<sup>-3</sup>) | Average pressure gradient<br>(Pa-m<sup>-1</sup>) |
| --- | --- | --- | --- | --- | --- |
| common carotid<br>sized | 0.03 | 0.006 | 0.0006 | $2.83 \times 10^6$ | 550.16 |
| aorta sized | 0.10 | 0.020 | 0.0025 | $6.11 \times 10^5$ | 998.32 |

The input parameters including the tube resistances and the pressure gradient set-up used in the full set of numerical experiments are illustrated in Table 1. The fluid properties are chosen to match blood (under the Newtonian fluid assumption) with the density and effective viscosity values as 1060 kg.m^3^ and 0.003 Pa.s respectively. The wall properties of the idealised cylindrical vessels, namely the vessel wall density and Poisson’s ratio were 1500 kg.m^3^ and 0.2 respectively, based on reported values in the literature^[47,21]^. For each simulation case, we compute the VDR *ϕ*_*v*_. Additionally, we compute a space-time averaged volumetric descriptor of dissipation defined using the following set of relations:

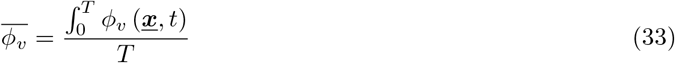

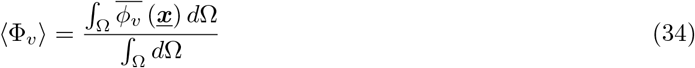

where Ω denotes the entire volume of the cylindrical tubes being considered; the first relation computes a time average over the pulse period *T* ; and the second relation computes a volume average of the time-averaged dissipation.

## 3 Viscous dissipation in aorta with left ventricular assist device

### 3.1 Computational modeling of hemodynamics driven by LVAD

For this study, a patient-specific arterial network comprising the aortic arch and branch arteries extending up to the iliofemoral arteries was obtained from Computed Tomography (CT) images, using 2D planar segmentation and lofting techniques implemented in the SimVascular software^[36]^. For model details refer to SimVascular^[42,36]^ and associated vascular model repository^[32]^, as well as our prior work^[29]^. The resulting 3D surface model represents the *baseline model* prior to LVAD outflow graft attachment. Subsequently, an image-based workflow was devised to attach a virtual cylindrical tube representing the LVAD outflow graft to the aortic arch in a manner that avoids intersection with nearby organs (heart, lungs) or bone boundaries (sternum, ribs). Extensive details of the workflow are provided in an earlier work^[33]^, and an outline is illustrated in Figure 2 panel a. Angle between LVAD outflow graft and the aorta was parameterized in terms of: (a) angle towards/away from aortic valve (referred to as *Inc* angles); and (b) angle towards left/right of the heart across the coronal plane (referred to as *Azi* angles). For this study we considered LVAD outflow grafts with 3 *Inc* angles: (1) perpendicular to aorta (*Inc90*), (2) 45 degree towards aortic valve (*Inc45*), (3) 45 degree away from aortic valve (*Inc135*); and 3 *Azi* angles: (1) 45 degree to the right of heart (*AziNeg45*), (2) perpendicular to coronal plane (*Azi0*), and (3) 45 degree to the left of heart (*Azi45*). Together, these comprise a set of 9 different LVAD surgical anastomosis models, as shown in Figure 2 panel b. Blood flow through each of these 9 LVAD graft anastomosis models, as well as the baseline model, was simulated using a stabilized finite element solver for incompressible Newtonian fluid mass and momentum balance as implemented in the SimVascular suite. The computational domain was discretized into linear tetrahedral elements with maximum edge size ≈ 0.67 mm as informed by prior mesh convergence studies using SimVascular for large artery hemodynamics (see for example^[23]^). Additional region based mesh sie refinements were conduced for those regions with small branching vessels, such as the abdominal aorta. Hemodynamics in the baseline model was driven by prescribing a physiologically measured pulsatile flow profile at the aortic root inlet. For the 9 LVAD anastomosis models, the aortic root inlet was assumed to be closed and modeled as a rigid wall. This assumption is based on existing studies documenting that the aortic valve remains continuously closed during periods of LVAD support^[19,26,40,8]^, and any trans-valvular flow is only intermittent contributing to a low level of flow^[25]^. For these LVAD models, hemodynamics was instead driven by prescribing a set of 3 different inflow profiles: (a) constant uniform flow over time; (b) flow with low extent of pulse modulation; and (c) flow with a high extent of pulse modulation, with the profiles for b. and c. adapted from prior studies^[16]^ (*see also Section 2.4)*. Time-averaged inflow was fixed at 4.9 L/min for all cases, with a total of 27 hemodynamics simulations across 3 inflow profiles, and 9 LVAD anastomosis models. We present the mean flow, flow ranges, and pulsatility index for the flow profiles considered in Table 3. Across all 27 simulations, boundary conditions at each outlet was kept fixed, and were assigned as 3-element Windkessel boundary conditions with resistance and compliance parameters obtained from existing literature^[32]^. Details on inflow and boundary conditions, and other numerical specifics, are provided in prior work^[33]^, and have also been included in Supplementary Material for brevity and conciseness of presentation.

**Figure 2:**
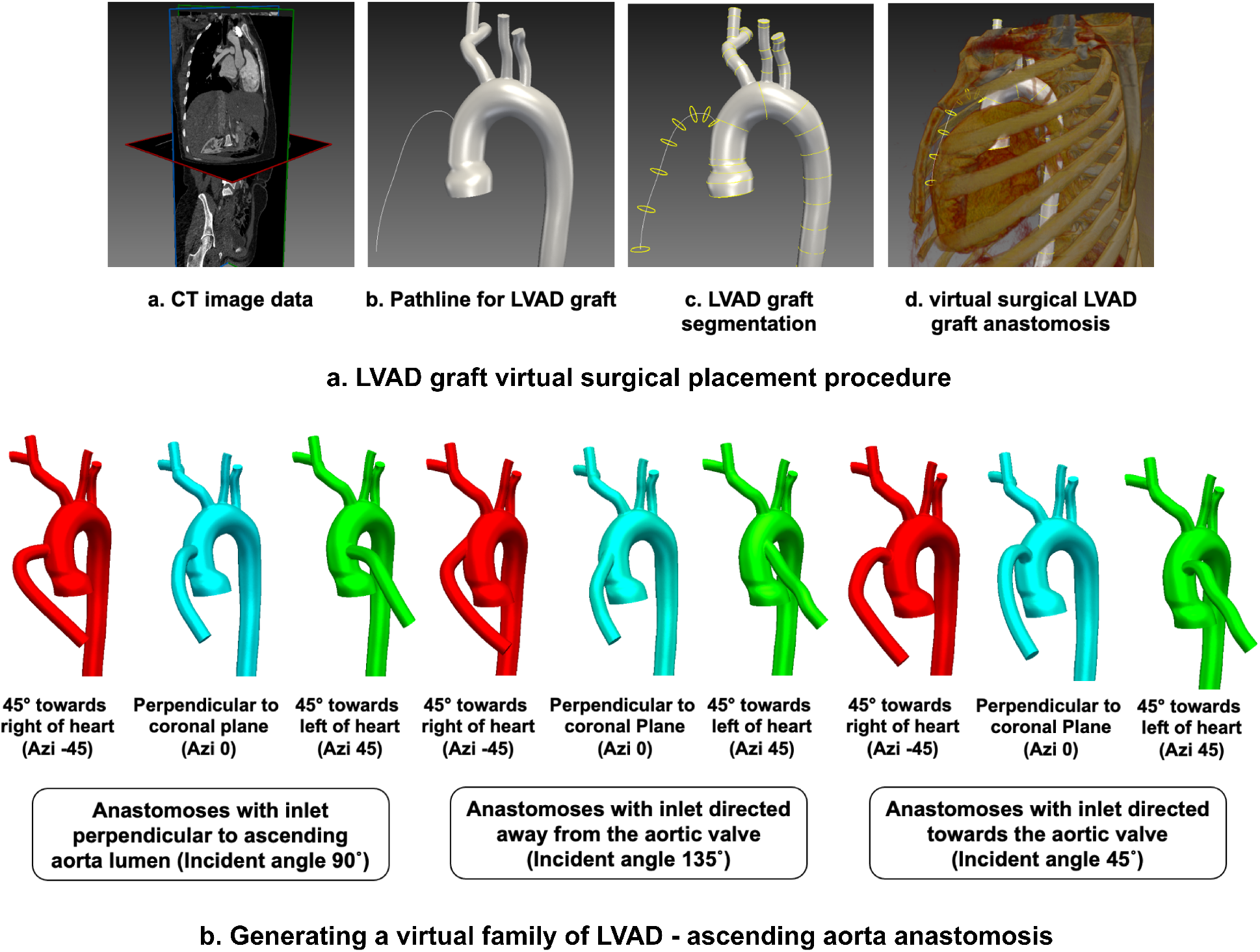
An illustration of LVAD modeling method as described in Section 3.1. Panel a. demonstrates the workflow for image-guided graft placement, using SimVascular image-processing toolkit. Panel b. depicts the 9 different graft anastomoses, created by varying graft attachment angles towards/away from the heart, and towards/away from the aortic valve.

### 3.2 Computing viscous dissipation rate based on CFD data

Computed blood flow velocity data from the third pulse cycle for each of the 27 LVAD cases, as well as the baseline case, were used to calculate the spatiotemporally varying VDR values using the same relation used in Section 2:

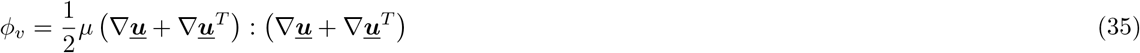

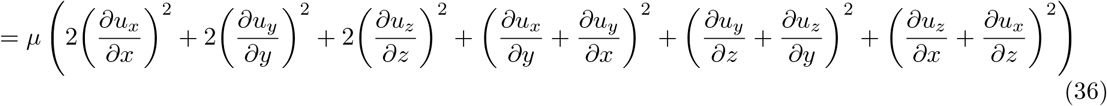

where the individual velocity gradient components were computed using built in discrete gradient filters in the open source VTK library^[34]^. Additionally, the space-time varying *ϕ*_*v*_ field data was further used to define a volume and time averaged VDR descriptor ⟨Φ_*v*_⟩ as defined in Section 2.4, but modified as follows. First, the time averaged value of *ϕ*_*v*_ is computed as:

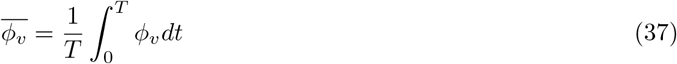

where the cycle average is obtained over the third pulse cycle of simulation time. Next, a volume average of 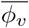 was computed over the aortic arch (after removing the contribution from LVAD outflow graft) and the abdominal aorta (after removing the contribution from the renal and mesenteric branching vessels) as follows:

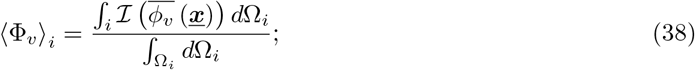

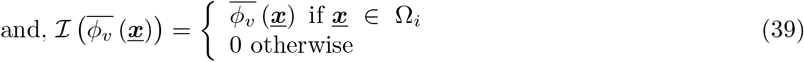

where Ω_*i*_ denotes respectively the domain for the aortic arch and abdominal aorta, and ℐ is an indicator function isolating the volume flow data in the respective domains. The VDR for total flow *ϕ*_*v*_ as well as the averaged dissipation descriptor ⟨Φ_*v*_⟩ were computed for all 27 LVAD flow scenarios considered, as well as for the baseline aorta without an LVAD.

## 4 Results

### 4.1 Parametric analysis of dissipation in idealized cylindrical tubes

The averaged dissipation metric ⟨Φ_*v*_⟩ as defined in Section 2.4, was computed for each of the 68 total parametric combinations considered for the cylindrical tube equivalent in diameter to the common carotid artery (as outlined in Figure 1). The resulting ⟨Φ_*v*_⟩ values are illustrated for all combinations in Figure 3. Panel a. depicts the values of ⟨Φ_*v*_⟩ computed based on the total flow velocity, while panel b. depicts the same for oscillatory components of the velocity only. Likewise, the computed ⟨Φ_*v*_⟩ values for all 16 parametric combinations considered for the cylindrical tube equivalent in diameter to the aorta (see Figure 1) are illustrated in Figure 4. Similar to Figure 3, panel a. depicts the dissipation metric computed based on the total velocity, while panel b. presents the same computed for oscillatory components of the velocity only. Observations from both Figure 3 and 4, indicate that the mean or zero-frequency contribution to flow dominates the total viscous dissipation. For the carotid equivalent tube, we observe that higher frequency contribution in the flow leads to lower extent of dissipation. We also observe that pulsatility index influences total dissipation as well as oscillatory contribution to dissipation, where for the same frequency and same mean flow, higher amplitude of pulsation leads to higher dissipation. The changes in total dissipation with pulsatility index is low (due to dominance of mean flow contribution), however the trends can be clearly seen when oscillatory flow contributions are considered as shown in Figure 3, panel b. Observations for the aorta equivalent tube also indicate same trends - higher pulsatility, and lower frequency contributions, lead to higher extent of dissipation. Since the pulse profiles used in the parametric simulations for the aorta equivalent tube are combinations of frequencies, it is important to note that across the low modulation, high modulation, and healthy aortic pulse profiles the contribution of the lower frequency components increase (see also Figure 1 panel c.). Additionally, for all cases considered we observe very small percentage changes in the averaged dissipation metric Φ_*v*_ with tube wall elasticity values; except for the scenario with completely rigid walls. Generally, across the 84 total scenarios considered for both tube diameters, the rigid tube has lesser extent of dissipation when considering the total flow (mean + oscillatory), in comparison to elastic tubes. This relatively low influence of wall elasticity, when compared to frequency and pulsation, can be explained by further demonstrating the variation of computed VDR *ϕ*_*v*_ for the elasticity ranges considered, as illustrated in Figure 5. The figure indicates that across physiologically realistic elasticity values as considered here, for frequencies that dominate the flow profiles (1-5 Hz regime), the resultant VDR varies very slowly as function of wall elasticity. We further note that, the theoretical estimates for a perfectly rigid walled tube (see Section 2.2), are different from those of finite elasticity tubes with a very high elasticity value (see Section 2.3). Theoretically, the presence of a finite elasticity will enable flow to move more freely than in perfectly rigid walled tube (as noted in^[46]^), which can enable flow-rates in finite elastic tubes to peak higher than in rigid tubes for same pulsation. This inference is further supported by the computed flow rate ranges for the cylindrical tube cases as reported in Table 2.

**Table 2:** A comparison of outflow rate ranges and pulsatility indices for a rigid walled (E inf.) vs an elastic walled (E = 1MPa) vessel, computed as a result of numerical experiments on idealised aorta-sized cylindrical vessel. Higher peak outflow rates and pulsatility indices were observed for the elastic walled tube as compared to the rigid walled tube.

| **Elasticity** | **Pulse profile** | **Mean Flow**<br>**(L.min<sup>-1</sup>)** | **Range**<br>**(L.min<sup>-1</sup>)** | **Pulsatility**<br>**index** |
| --- | --- | --- | --- | --- |
| **Rigid**<br>**tube**<br>($E \text{ inf.}$) | low modulation | 4.90 | 4.77 - 5.02 | 0.05 |
|  | high modulation | 4.90 | 4.90 - 5.17 | 0.11 |
|  | aortic | 4.91 | 4.31 - 5.43 | 0.23 |
| **Elastic**<br>**tube**<br>($E = 1MPa$) | low modulation | 4.91 | 4.27 - 5.96 | 0.34 |
|  | high modulation | 4.91 | 4.24 - 6.09 | 0.38 |
|  | aortic | 4.91 | 4.45 - 6.30 | 0.38 |

**Table 3:** Characteristics of the pulse-modulated LVAD inflow profiles and baseline aortic inflow profile used in the aortic hemodynamics study. Note that baseline aortic flow has a small negative flow-rate value due to effects of diffuse wave reflections along the arterial tree when measured at a location away from the aortic valve.

| **Pulse profile** | **Mean flow (l/min)** | **Range (l/min)** | **Pulsatility Index (PI)** |
| --- | --- | --- | --- |
| **low modulation** | 4.90 | 3.0 - 7.0 | 0.81 |
| **high modulation** | 4.90 | 1.0 - 10.0 | 1.82 |
| **aortic** | 4.90 | -0.6 - 18.0 | 3.83 |

**Figure 3:**
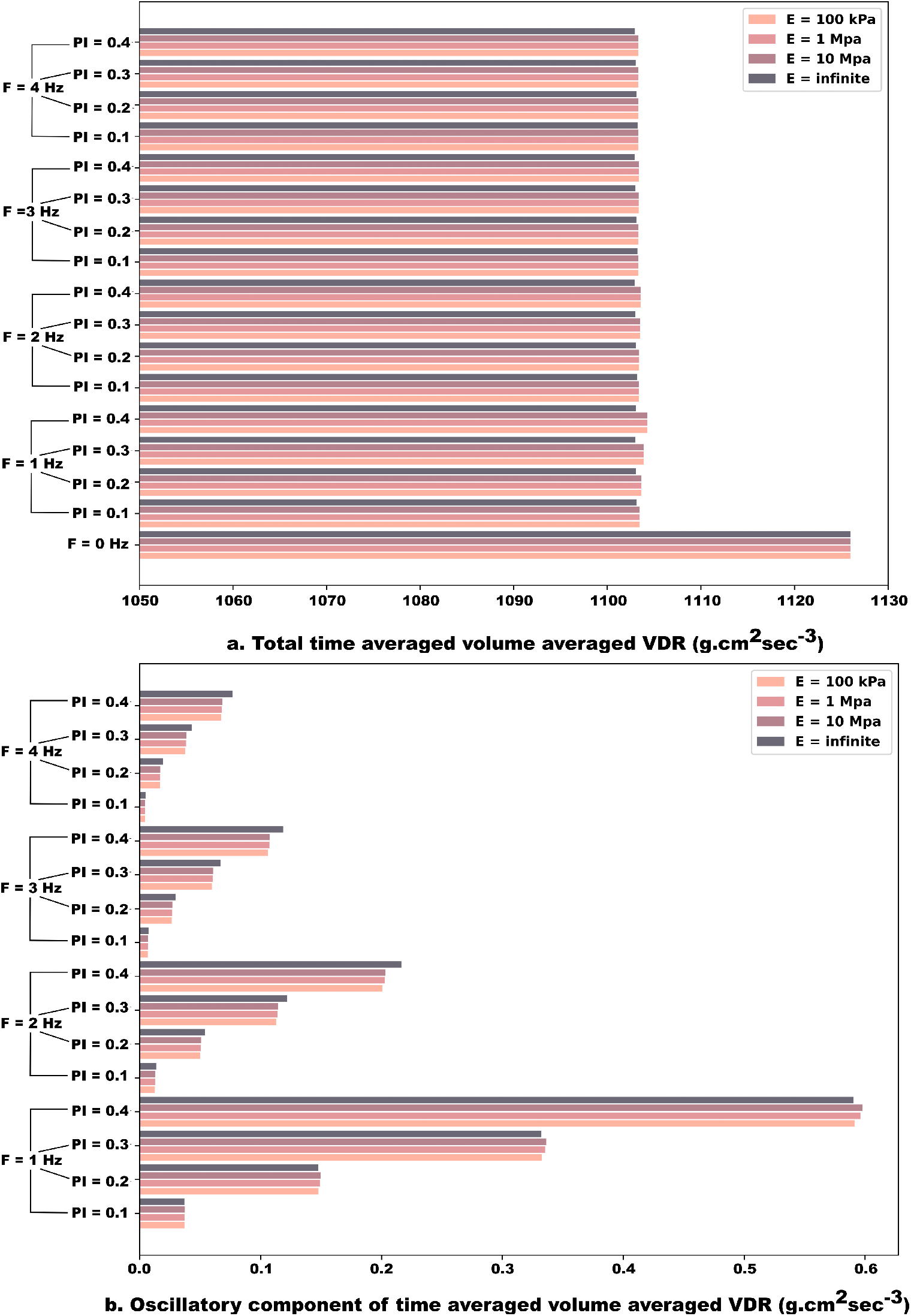
Computed viscous dissipation rate (VDR) for pulsatile flow through an idealized cylindrical vessel equivalent to the Common Carotid Artery (CCA) size, for a set of 16 pressure gradient pulses and a constant pressure gradient inflow. Panel a. depicts the total (steady + oscillatory contribution) volume integral of time averaged VDR in CCA sized cylindrical tube. Panel b. depicts the oscillatory velocity component contribution to volume integral of time averaged VDR in CCA sized cylindrical tube.

**Figure 4:**
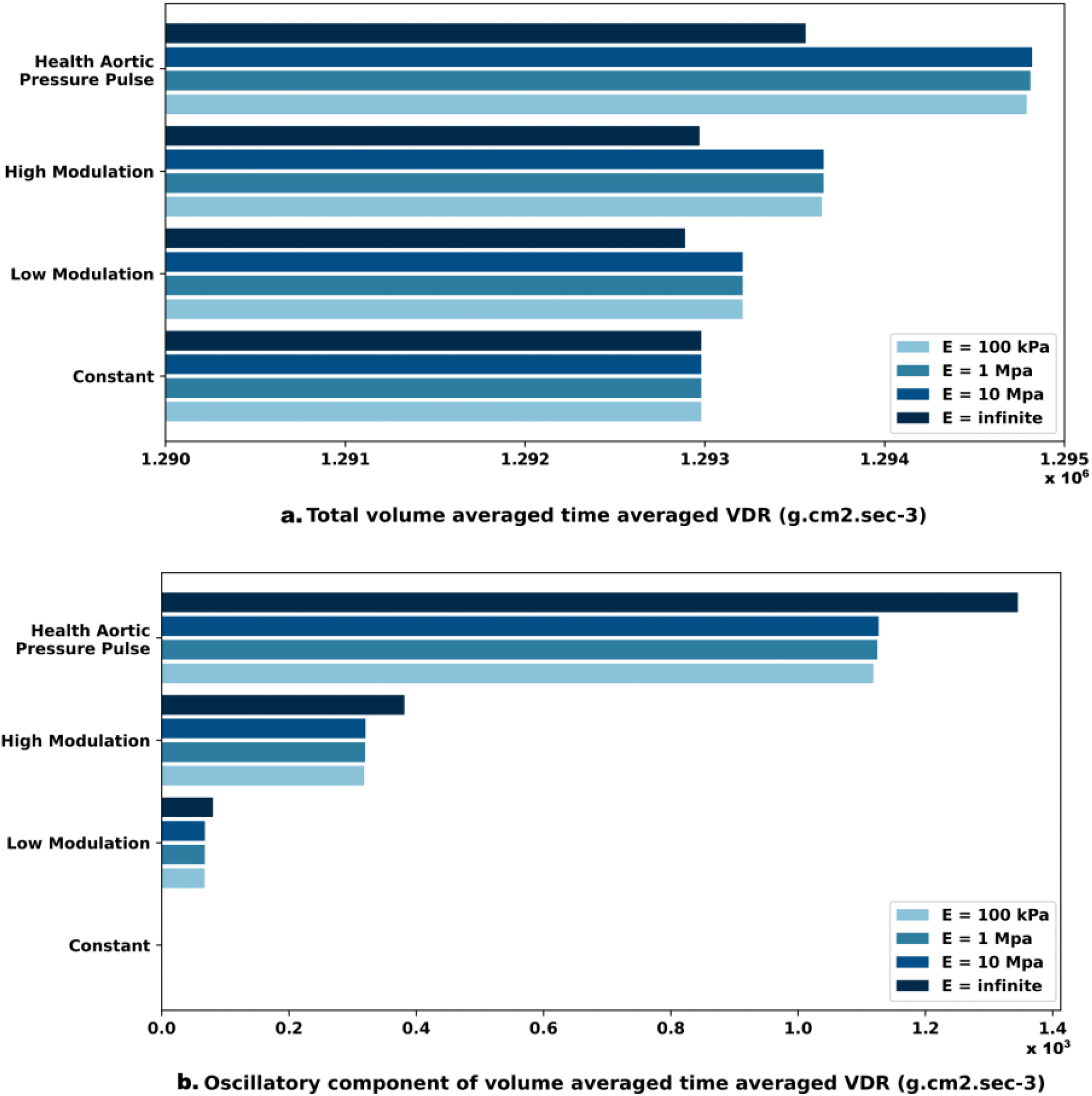
Computed viscous dissipation rate (VDR) for pulsatile flow through an idealised cylindrical vessel equivalent to the Aorta size, for a set of four pressure gradient pulses. Panel a. depicts the total volume integral (steady + oscillatory contribution) of time averaged VDR in aorta sized cylindrical tube. Panel b. depicts the oscillatory velocity component contribution to volume integral of time averaged VDR in aorta sized cylindrical tube.

**Figure 5:**
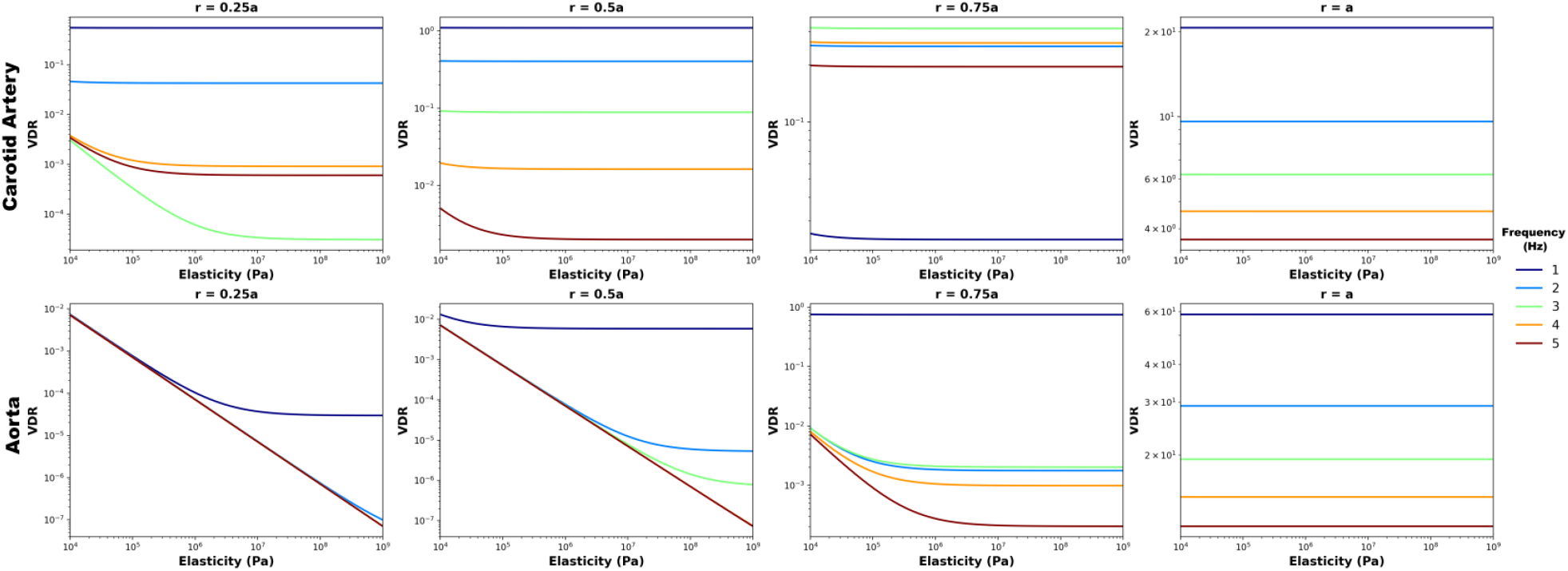
Plot depicting variation in Viscous Dissipation Rate (VDR) with wall elasticity (E) for varying inflow pressure gradient pulse frequencies (Hz) in idealised cylindrical tubes of sizes equivalent to a. common carotid artery (CCA) and b. aorta. The VDR variation with E is plotted along points located at a radius ‘r’ from the tube center-line.

### 4.2 Viscous dissipation analysis in aorta with an LVAD

Viscous dissipation *ϕ*_*v*_ and the averaged dissipation descriptor ⟨Φ_*v*_⟩ as defined in Section 3.2 for each of the 27 LVAD flow scenarios considered are illustrated in Figure 6 for the aortic arch region, and in Figure 7 for the abdominal aorta region. Specifically, panel a. in Figure 6 depicts isosurfaces of computed VDR *ϕ*_*v*_for the aortic arch region (LVAD outflow graft and branch vessels included), for each of the 27 LVAD flow scenarios. Each row in panel a. denotes the outflow graft orientation towards/away from aortic valve (*Inc angles*); and columns denote pulse modulation categorized per graft orientation with reference to the coronal plane (*Azi angles*) as detailed in Section 3.1. Panel b. in Figure 6 depicts the ratio between the computed averaged dissipation descriptor ⟨Φ_*v*_⟩ for the aortic arch region Ω_*i*_ for each LVAD configuration, as compared against corresponding baseline flow. Likewise, panels a. and b. for Figure 7 respectively represent the corresponding illustrations for *ϕ*_*v*_ isosurfaces, and ratio of ⟨Φ_*v*_⟩ between LVAD scenarios and baseline flow, for the abdominal aorta region. The isosurface plots for the various LVAD cases illustrate the influence of the impingement of the LVAD outflow jet in the aortic arch region. We observe that aortic hemodynamics originating from this jet impingement, depends upon the outflow grafts as well as the extent of pulsation. Specifically, while outflow graft angles govern the orientation of the LVAD outflow jet and the location of jet impingement along the aorta wall; the extent of pulse modulation influences the intensity of the impingement. Additionally, regions of stasis in the aortic arch at the root of proximal aorta are indicated in Figure 6 panel a. as regions of low to no VDR emanating from slow arrested flow. These regions are highly prone to thrombus formation driven by flow stagnation, which further relates to risk of stroke and other complications; providing thereby a link between VDR-based descriptors and pathological complications. As the flow generated by the jet impingement rolls along the aorta into the abdominal aorta, we see similarly in Figure 7, panel a. that the graft angles and pulsation continue to influence extent of dissipation in the abdominal aorta, altough the extent of dissipation is significantly lower. Observations from Figure 6, panel b. indicate that the averaged extent of dissipation (quantified here by ⟨Φ_*v*_⟩ values) in the aortic arch is higher compared to the baseline flow for each of the 27 different LVAD flow scenarios considered (that is, ratios are all greater than unity). The computed ratios are strongly influenced by the graft angle towards/away from the valve (*Inc angles)* - decreasing in value across *Inc45, Inc90*, and *Inc135* cases. We also observe that, consistently across all LVAD graft angles considered, extent of dissipation in the aorta increases with increasing pulse modulation. On the contrary, observations from Figure 7, panel b. indicate that the averaged extent of dissipation in the abdominal aorta is lower compared to the baseline flow for each of the 27 different LVAD flow scenarios considered (that is, ratios are all lesser than unity). We also note that, similar to aortic arch, extent of dissipation in the abdominal aorta still increase with increasing pulse modulation for all LVAD graft angles considered. However, unlike the arch region, dissipation in the abdominal aorta is more prominently influenced by the outflow graft angle towards the left/right of the heart (*Azi angles*). These observations are in alignment with vorticity generation at LVAD jet impingement location, and vorticity transport and dissipation into descending and abdominal aorta post impingement, as illustrated in detail in our prior work^[33]^, hinting at the central role of jet impingement phenomena in determining energetics in hemodynamics during LVAD operation.

**Figure 6:**
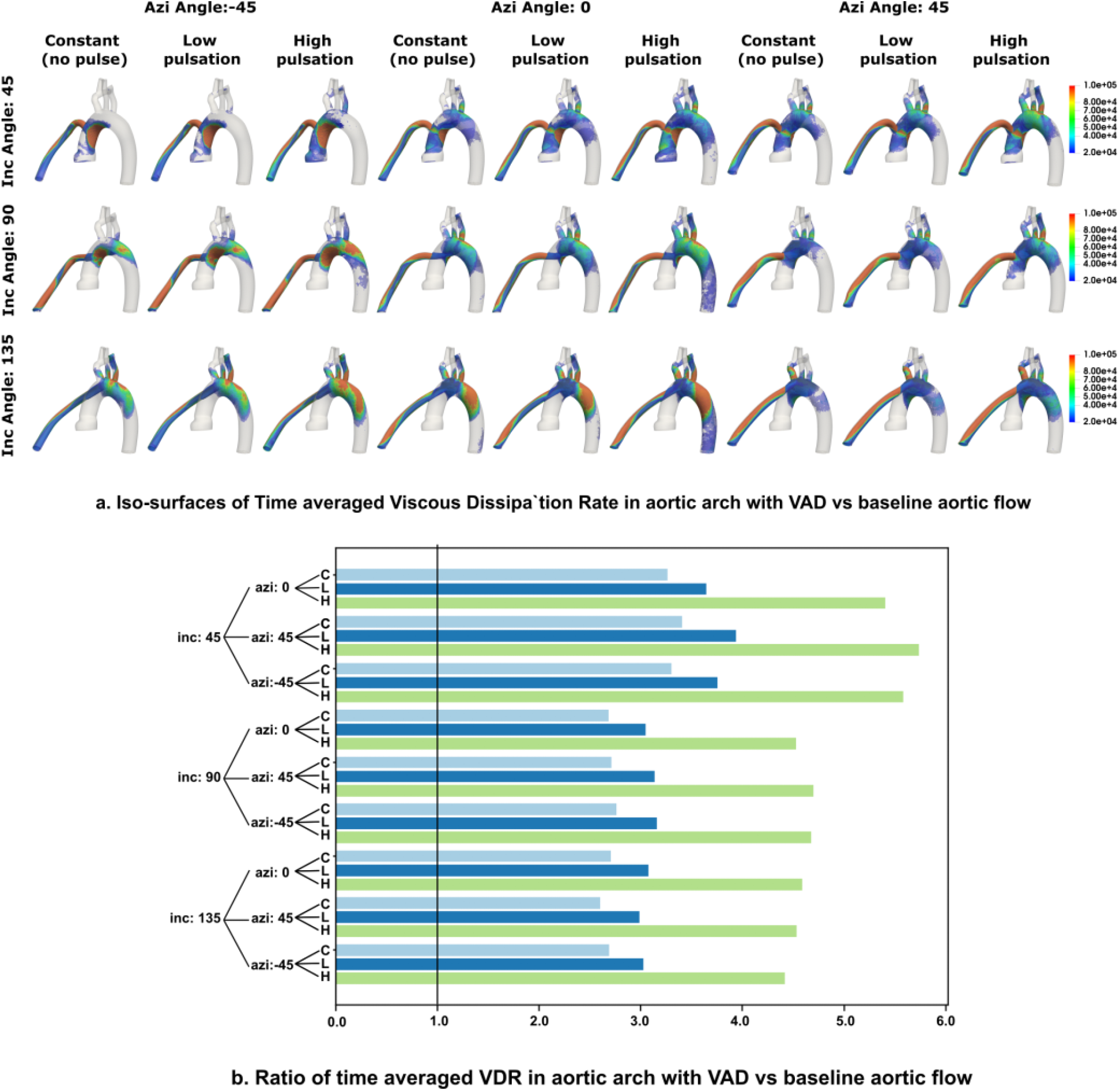
Simulated viscous dissipation quantifier for the aortic arch region for all 27 LVAD flow scenarios. Panel a. presents the time averaged viscous dissipation rate (VDR) iso-surfaces, panel b. presents the ratio of the volume averaged VDR metric for the aortic arch compared to the corresponding baseline arch. Ratio = 1.0, marked on panel b. represents baseline flow without LVAD. The inflow pulse profiles are depicted by C (constant flow), L(low pulsation) and H(high pulsation).

**Figure 7:**
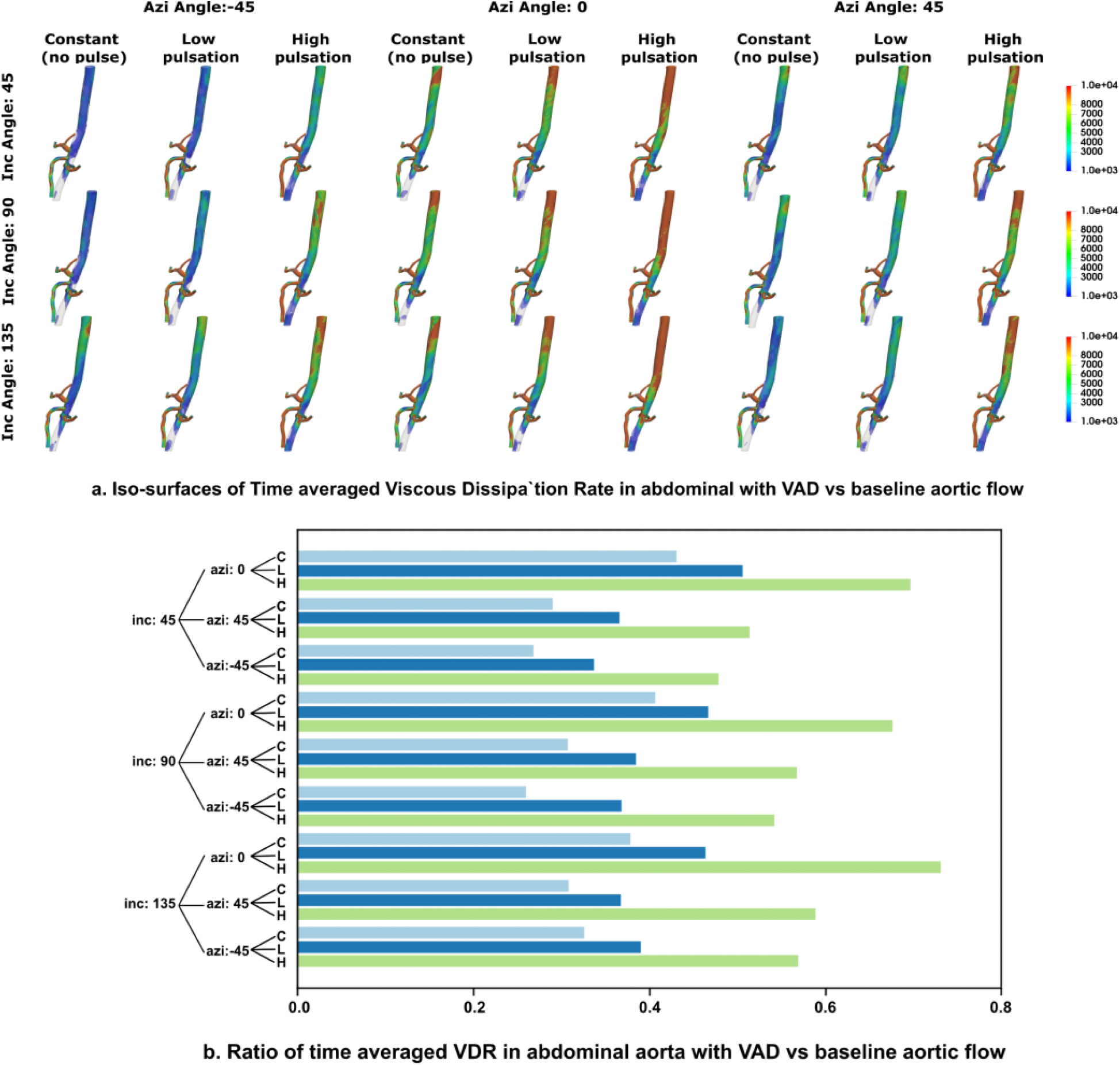
Simulated viscous dissipation quantifier for the abdominal aorta region for all 27 LVAD scenarios. Panel a. presents the time averaged viscous dissipation rate (VDR) iso-surfaces, panel b. presents the ratio of the volume averaged VDR metric for the abdominal aorta region as compared to the corresponding baseline abdominal aorta region. Ratio = 1.0, marked on panel b. represents baseline flow without LVAD. The inflow pulse profiles are depicted by C (constant flow), L(low pulsation) and H(high pulsation).

## 5 Discussion

Here we conducted a systematic multi-parameter analysis of viscous dissipation for: (a) flow with varying pulsatility indices and frequency in cylindrical tube of diameter equivalent to the common carotid artery; (b) flow with varying pulse profiles (over time) in cylindrical tube of diameter equivalent to the aorta; and (c) patient-specific vascular model with an attached LVAD outflow graft driving flow with varying pulse profiles (same profiles selected as in case b). The results from a total of 112 different simulation cases elucidate how energy dissipation in arterial hemodynamics in the context of mechanical circulation support is determined through an interplay of surgical parameters (graft angles), flow pulsation, and vessel wall properties. The single vessel simulations, based on analytical expressions, illustrated the effects of pulsation and wall elasticity, without considering details of vascular anatomy and surgical anastomoses. The patient-specific study demonstrated the effects of pulsation, anatomy, and surgical anastomoses; and while the vessel walls were assumed to be rigid, compliance effects due to elasticity were incorporated through downstream boundary conditions. Thus, the single vessel and the patient-specific simulations were used as complementary analyses to understand underlying factors governing energy dissipation. Our findings suggest that for physiologically relevant LVAD MCS scenarios, wall elasticity plays a potentially second order role when compared to the strong influence that pulsation and surgical graft angles play in determining energy dissipation. Pulsation and frequency contributions by itself influence the extent of energy dissipation as indicated in the single vessel simulations. When viewed in conjunction with the LVAD graft angles in anatomically realistic vasculature, the influence of pulsation becomes more pronounced, as the LVAD jet impingement drives the aortic hemodynamics. Furthermore, for the patient-specific model, the comparison of viscous dissipation for the various LVAD cases against that for baseline flow, illustrates how the LVAD outflow jet impingement leads to an altered state of hemodynamics in the aorta when compared to the baseline. Specifically, this can be interpreted based on a two jet flow model for LVAD hemodynamics, as proposed in our prior work^[33]^. The baseline aortic hemodynamics is driven by the “*aortic jet*” emanating from aortic valve opening during ventricular systole. However, during LVAD operation, aortic hemodynamics is driven by the “*LVAD outflow jet*” which traverses across the aorta centerline and impinges on the aorta wall. In prior work^[33]^ we developed this two jet flow explanation and demonstrated how hemodynamics driven by the LVAD outflow jet differs from that driven by the baseline aortic jet in terms of the state of flow, mixing, and wall shear. Here, we further advance this understanding by showing how the flow driven by the two jets differ in terms of the extent of hemodynamic energy dissipation.

Findings from this study further advance our understanding of altered state of aortic flow during LVAD operation, with a focus on viscous energy dissipation. We identify three significant aspects from the study. First, we explore in detail the fundamental interplay of frequency, pulsation, and wall elasticity, in determining state of energy dissipation in arterial hemodynamic scenarios. The availability of simplified expressions as outlined in Section 2 enables us to explore the effects of wall properties, without directly conducting substantially more expensive fluid-structure interaction simulations. Wall elasticity is related to vessel wall stiffening, which in itself is a consequence of aging, or other vascular pathologies. Frequency and pulsation have emerged as critical factors for consideration in LVAD design and operation, and our study provides some additional insights into the role of pulse-modulation in LVAD therapy as outlined in other works^[31,43]^. We have ensured that the parametric analysis of VDR due to anastomosis and pulsation remained decoupled from specific pump hardware features such as blade sweep and pump part design. This enables us to discuss the effects of the fundamental variables considered here in a device/manufacturer neutral manner. It was not our goal to tie this analysis to a specific manufactured pump design. Second, energy dissipation has previously been used to develop hemodynamic descriptors and indices for flow efficiency assessment in reconstructive surgical procedures such as Glenn and Fontan for congenital heart disease patients^[10]^. Our findings establish that energy dissipation can also be similarly applied to devise flow efficiency indices in patients on LVAD support as well. As a specific example, we have discussed the potential role of energy dissipation as an additional indicator for arrested flow and thrombogenicity in Section 4.2. As another potential example, the relation between energy dissipation and efficiency of perfusion in distal organ beds away from aortic root can serve as additional clinical variable for treatment efficacy assessment. These aspects need further exploration in future efforts. Third, our patient-specific analysis incorporated varying graft surgical parameters, and pulse modulation; and compared the VAD-driven flow to an estimate of the baseline flow without an LVAD. This methodology enabled quantifying the extent by which the LVAD jet driven flow differs from baseline; which is information that may not be otherwise available or generated. There is increasing interest in clinical community on what physiological flow features are adversely altered during LVAD operations^[31]^, and our study provides the foundations to develop relevant hemodynamic descriptors for this altered flow state for improved understanding of etiology of post-surgical complications in VAD therapy. Flow imaging is conventionally done using techniques like 4D flow MRI, which is not suitable for LVAD. Hence, a comprehensive in silico parametric approach as outlined here based on fundamental flow physics theories provides a suitable avenue for advanced hemodynamic assessment for patients on LVAD support. Each of these aspects could further inform optimization of LVAD outflow graft surgical anastomosis in conjunction with stroke and bleeding risk estimates and other hemodynamic parameters - thereby laying the basis for future discussions on utilizing hemodynamic energy dissipation for evaluating LVAD surgical outcomes and efficiency.

The computational analysis presented here was based on several key underlying assumptions, with associated limitations. First, we compared the extent of dissipation using an averaged viscous dissipation metric Φ_*v*_ for all of our simulations. This was employed as a single aggregate quantifier that can differentiate the spatiotemporal complexity of energy dissipation in flow for different LVAD cases, and not as a means to compare point-wise energy dissipation for varying parameters. Similar reduced order quantifiers for energy dissipation can also be obtained using Lagrangian approaches by integrating VDR along trajectories of flow tracers. This has not been explored here, but remains an aspect of continued interest. Second, here we have not considered any turbulence modeling, which may need to be considered for flow transition into turbulence locally in the jet impingement zone. Turbulence modeling using an additional eddy viscosity will lead to greater extent of viscous dissipation, making the numbers from our study more conservative estimates of dissipation. However, we anticipate that the overall parametric influence trends will remain similar even considering local turbulence. Third, here we have not considered ventricular effects of any kind, to focus simply on the surgical parameters and hemodynamics alone. For example, in the patient-specific case, we assume that the valve was always shut. As mentioned earlier, any additional flow due to intermittent valve opening, will reduce stasis in the aortic root, and add more dissipation to the baseline conservative estimates. Just like turbulence, we anticipate our parametric trends to remain similar. Additionally, the global pumping efficiency for LVADs has two major components: the energy dissipation contribution from the assisting the ventricular load and the dissipation due to flow in the arteries. In our study we have only focused on quantifying the latter. To emphasize this point, the overall energy requirements due to an increased ventricular load due in an advanced HF patient may additionally alter the energy dissipation rates in a systemic manner and thus energy requirements for blood flow circulation.

## 6 Concluding remarks

In summary, we present a comprehensive characterization of viscous energy dissipation in arterial hemodynamics, with application to circulatory support using LVADs. We conducted a complementary set of analyses using pulsatile flow in idealized cylindrical tubes, and patient-specific models with varying LVAD outflow graft angles and pulse-modulation. The findings illustrate the dominant effect of frequency, pulsation, and surgical attachment angles in determining state of energy dissipation, while a weaker influence of wall elasticity for LVAD relevant scenarios. The findings further advance our understanding of the central role played by LVAD jet impingement in determining how hemodynamics driven by the LVAD can differ from baseline physiological scenarios. The resulting comprehensive characterization of hemodynamic energy transport and dissipation can help devise innovative avenues to address a growing interest in improving LVAD therapy outcomes, and optimize the LVAD surgical configurations.

## Supporting information

Supplementary Video: VDR-combined-anim.mp4

## Data Availability

All data produced in the present work are contained in the manuscript. Human vascular model data obtained from open source Vascular Model repository (www.vascularmodel.com) associated with the open source SimVascular project (simvascular.github.io)

## Acknowledgements

This work was supported by a University of Colorado Anschutz-Boulder (AB) Nexus Research Collaboration Grant awarded to authors DM and JP. This work utilized resources from the University of Colorado Boulder Research Computing Group, which is supported by the National Science Foundation (awards ACI-1532235 and ACI-1532236), the University of Colorado Boulder, and Colorado State University. AS conducted the modeling, simulation, and quantitative analysis and post-processing for the study; and developed the computer codes and scripts for the single vessel analysis. KC conducted post-processing and synthesis of simulation data from single vessel study. DM designed the study, conducted data analysis and interpretation, and drafted the manuscript in collaboration with AS, EM, and JP. JP and EM guided the study design, and hemodynamic parameter selection alongwith clinical interpretation of the data. All five authors reviewed and finalized the draft, and are in agreement regarding the final content of the manuscript.

## Supplementary Material Information

### Animation of viscous dissipation patterns in aortic arch with LVAD

Alongwith the manuscript we have included a separate animation file named “*vdr-combined-anim*.*mp4*”, which presents a combined animation of viscous dissipation in the aortic arch for all 27 LVAD scenarios considered in this study. This animation provides a clear view of the dynamics of LVAD jet impingement onto the aorta wall and subsequent influence on viscous dissipation. The animation is 10 seconds long, and considering the pulse cycle as 1 second, the animation is thus rendered 10 times slower than actual pulse cycle.

### Additional visualization of viscous dissipation data for patient-specific model

Here we present additional visualization of the viscous dissipation rate data for the patient-specific model considered in this study. First, to complement the viscous dissipation results shown in Section 4.2, we provide in Figure S1 a visualization of the time-averaged VDR *ϕ*_*v*_ isosurfaces for the baseline aorta model without an LVAD. Additionally, we supplement the analysis of averaged dissipation ratio ⟨Φ_*v*_⟩ for the LVAD models computed with reference to the baseline model, by presenting an additional comparison in Figure S2. In this comparison, we compute the ratio of ⟨Φ_*v*_⟩ between the aortic arch region and the abdominal aorta region for each of the 27 LVAD scenarios considered in this study, and compare the ratios against the same computed for the baseline model (indicated in red). We clearly observe that in baseline case, flow in the abdominal aorta region has greater extent of energy dissipation compared to the aortic arch. Conversely, for all 27 LVAD scenarios considered, flow in the aortic arch has a greater extent of energy dissipation compared to the abdominal aorta. This further complements our explanations of energy dissipation due to hemodynamics originating from LVAD outflow jet impingement on the aorta wall.

### Boundary conditions used for modeling aortic hemodynamics driven by LVAD

In conjunction with the methodology outlined in Section 3.1, and following the details presented in our prior work^[33]^, here we have included the details of all inflow and outflow boundary conditions for the computational simulations of hemodynamics driven by LVAD. First, in Figure S3, we present all three pulse modulated inflow profiles used for the LVAD hemodynamics, and compare it against the baseline inflow profile. Lastly, in Table S1, we present the proximal and distal resistances *R*_*p*_ and *R*_*d*_, as well as the compliance estimated *C* for all the 3-element Windkessel boundary conditions for each outlet in the model.

**Figure S1:**
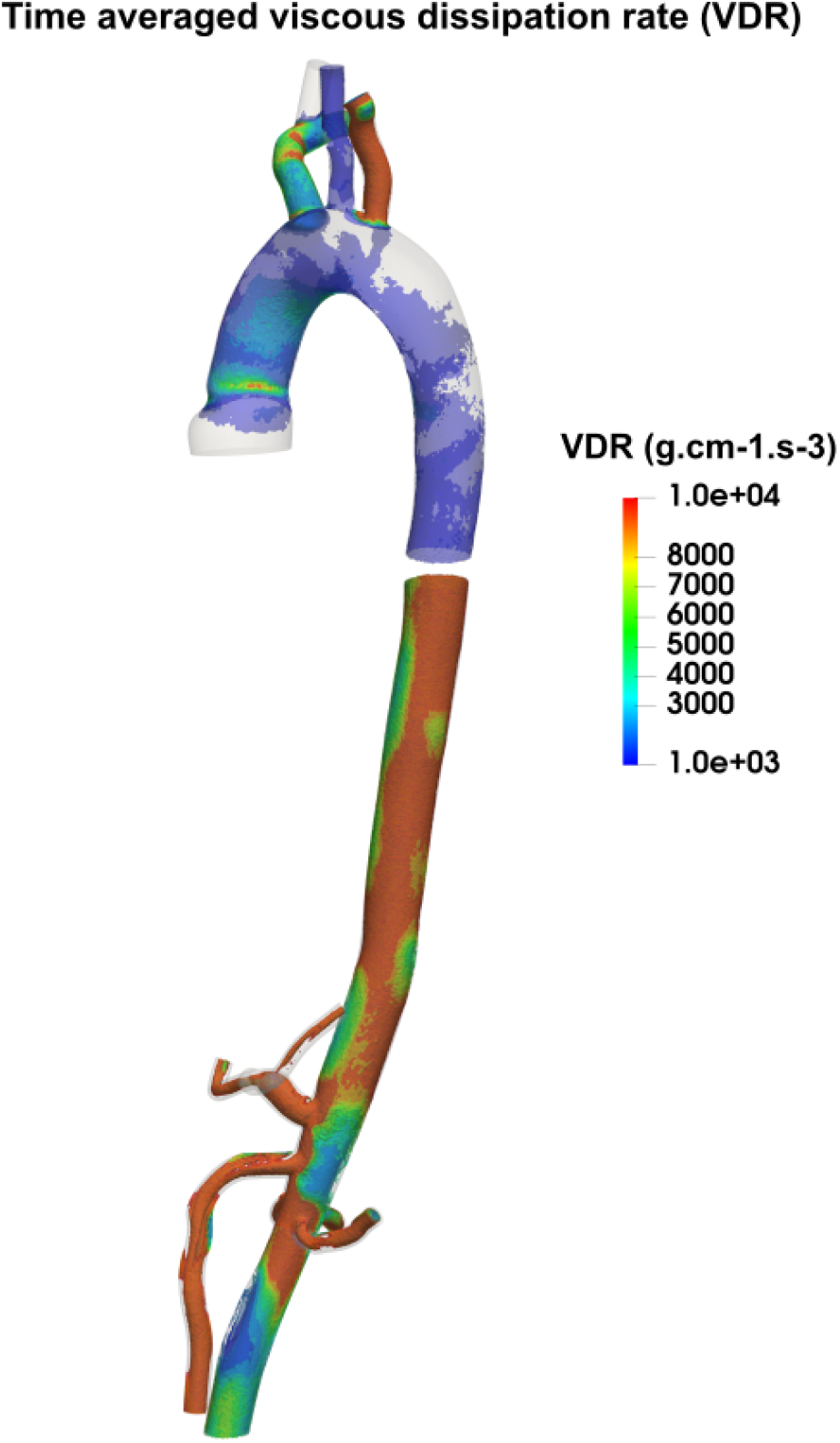
Iso-surfaces of time averaged Viscous Dissipation Rate (VDR) in the aorta without LVAD. Panel a. shows the aortic arch region and panel b. shows the abdominal aorta region.

**Figure S2:**
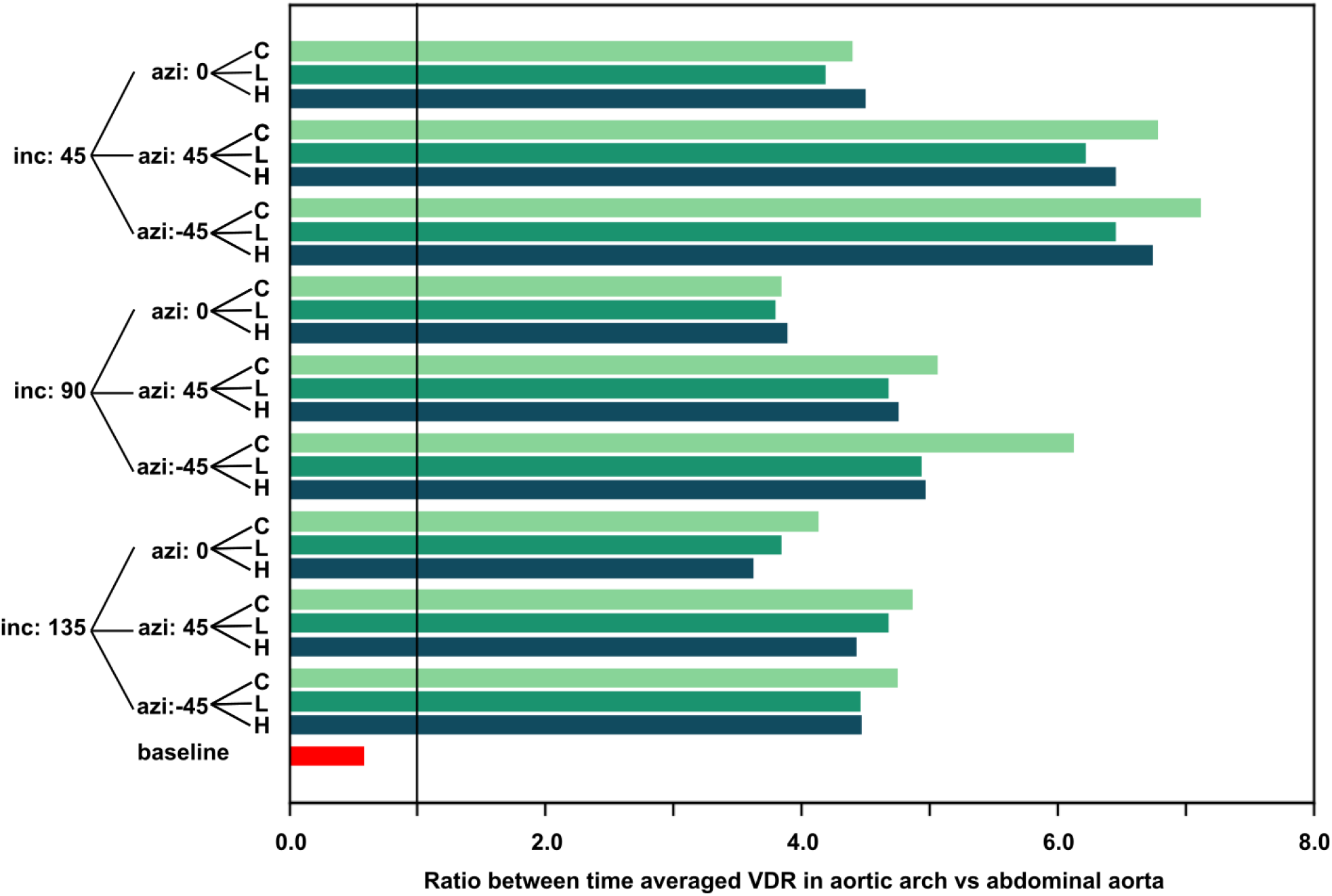
Ratios of computed Viscous Dissipation Rate descriptor for aortic arch vs abdominal aorta with VAD. Corresponding value for baseline is plotted in red for comparison. Note: the pulse modulations are depicted by C (constant flow), L(low pulsation) and H(high pulsation)

**Figure S3:**
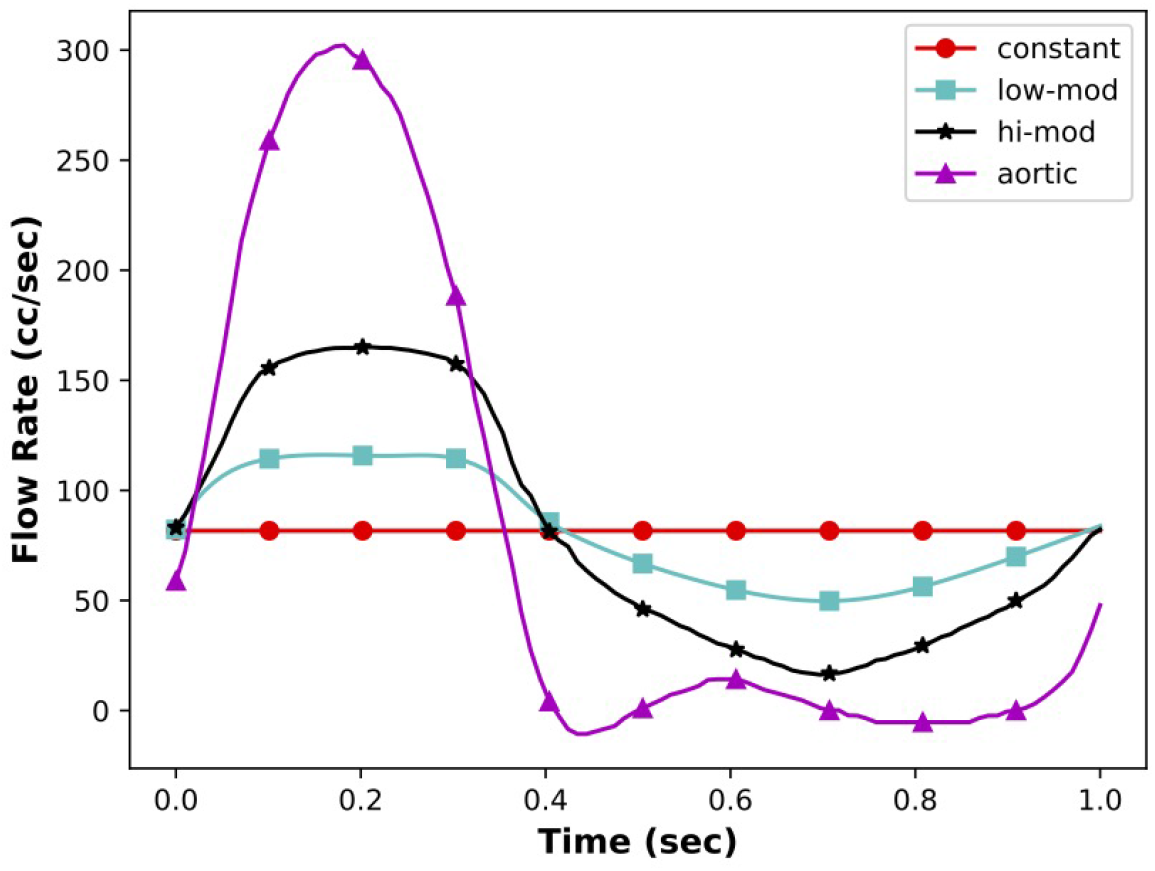
A plot depicting pulse modulated LVAD inflow profiles and baseline flow profile used in the aortic hemodynamics study.

**Table S1:**
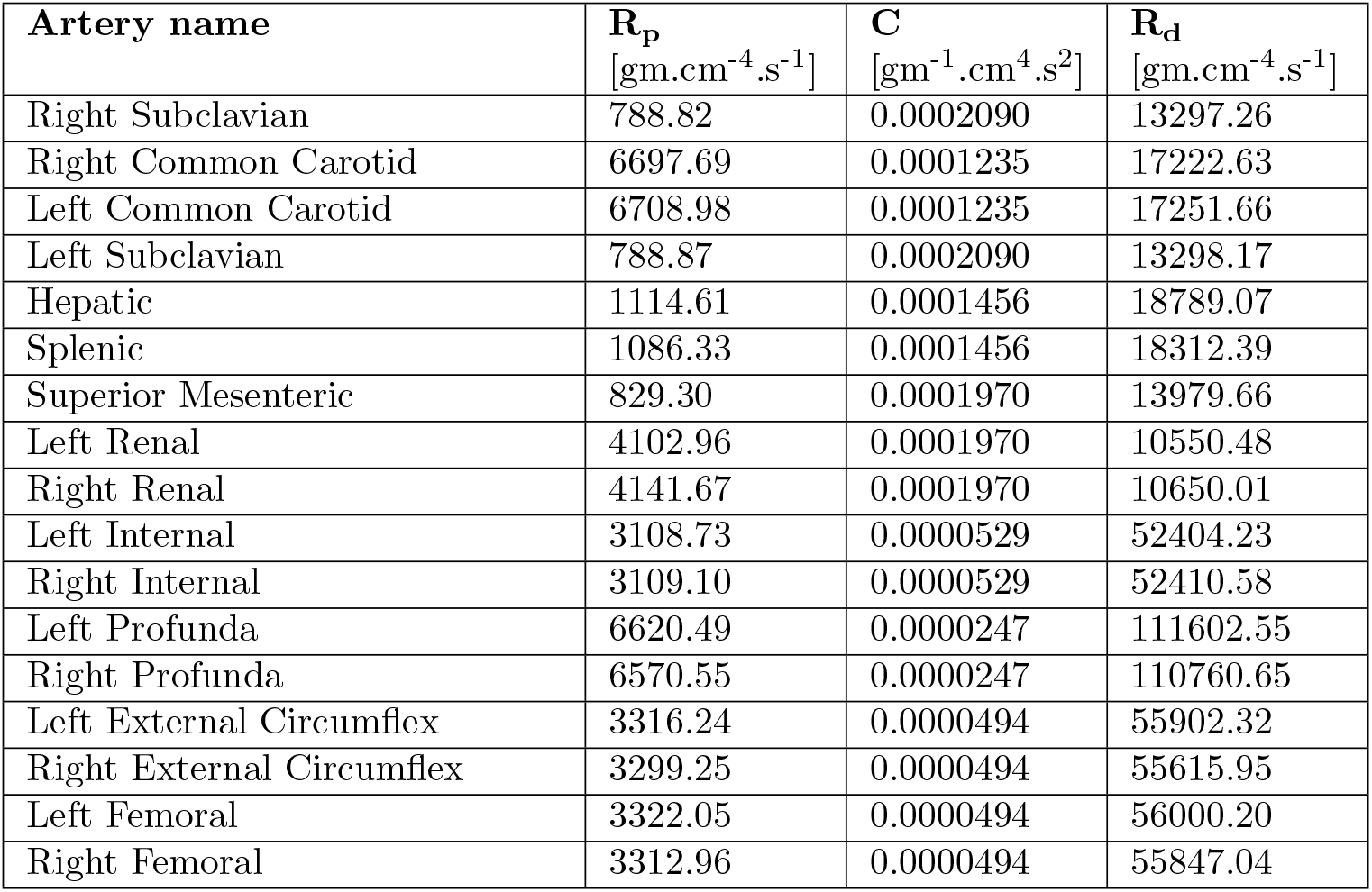
*A complete list of proximal resistance* **R**_**p**_, *capacitances* **C** *and distal resistance* **R**_**d**_ *values for three-element Windkessel boundary conditions assigned to all aorta model branch outlets*.

| Artery name | $\mathbf{R_p}$<br>[gm.cm<sup>-4</sup>.s<sup>-1</sup>] | $\mathbf{C}$<br>[gm<sup>-1</sup>.cm<sup>4</sup>.s<sup>2</sup>] | $\mathbf{R_d}$<br>[gm.cm<sup>-4</sup>.s<sup>-1</sup>] |
| --- | --- | --- | --- |
| Right Subclavian | 788.82 | 0.0002090 | 13297.26 |
| Right Common Carotid | 6697.69 | 0.0001235 | 17222.63 |
| Left Common Carotid | 6708.98 | 0.0001235 | 17251.66 |
| Left Subclavian | 788.87 | 0.0002090 | 13298.17 |
| Hepatic | 1114.61 | 0.0001456 | 18789.07 |
| Splenic | 1086.33 | 0.0001456 | 18312.39 |
| Superior Mesenteric | 829.30 | 0.0001970 | 13979.66 |
| Left Renal | 4102.96 | 0.0001970 | 10550.48 |
| Right Renal | 4141.67 | 0.0001970 | 10650.01 |
| Left Internal | 3108.73 | 0.0000529 | 52404.23 |
| Right Internal | 3109.10 | 0.0000529 | 52410.58 |
| Left Profunda | 6620.49 | 0.0000247 | 111602.55 |
| Right Profunda | 6570.55 | 0.0000247 | 110760.65 |
| Left External Circumflex | 3316.24 | 0.0000494 | 55902.32 |
| Right External Circumflex | 3299.25 | 0.0000494 | 55615.95 |
| Left Femoral | 3322.05 | 0.0000494 | 56000.20 |
| Right Femoral | 3312.96 | 0.0000494 | 55847.04 |

### Note on VDR expressions

A few additional details on the derivation and algebra of the expressions for VDR using analytical theories of pulsatile flows in rigid and elastic tubes developed in prior works^[46,45]^ are included here to supplement the discussion in Section 2 in the manuscript. We reproduce the expression for viscous dissipation:

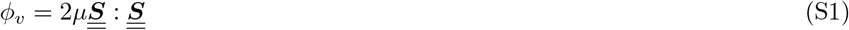

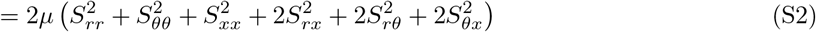

For cylindrical polar coordinates, using the definition of the vector calculus operators with the appropriate curvilinear coordinate derivatives, the strain-rate tensor components for velocity field (*u*_*r*_, *u*_*θ*_, *u*_*x*_) can be written out as follows:

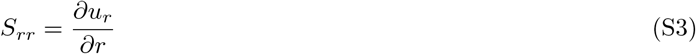

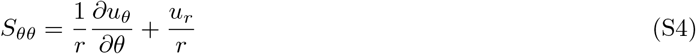

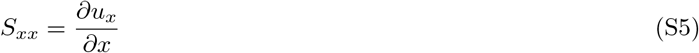

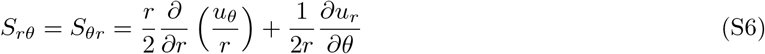

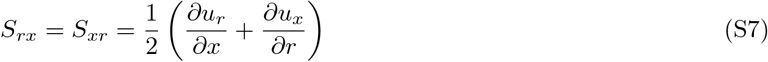

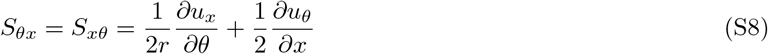

#### Pulsatile flow in rigid tubes

For the case of pulsatile flow in rigid cylindrical tubes, the generalized expression for the flow velocity, including both the steady and pulsatile contributions is written as:

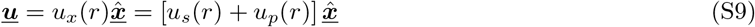

Based on this functional form of the velocity, the only non-zero contribution from the strain-rate tensor is:

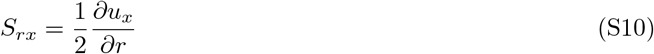

which leads to the following viscous dissipation rate:

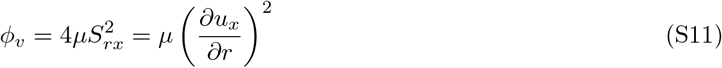

Using the decomposition into a steady and a pulsatile contribution:

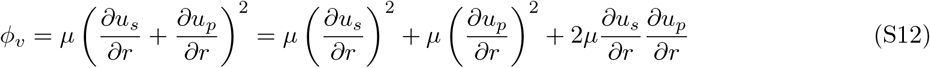

With the expressions for the steady and oscillatory contribution of velocity as outlined in Section 2.2 now we can obtain a series of final expressions for the various components of the viscous dissipation rate:

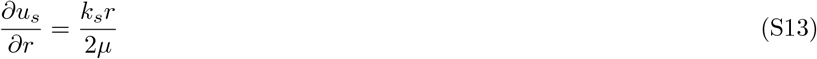

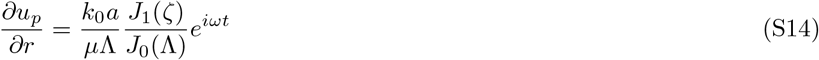

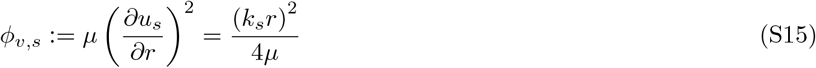

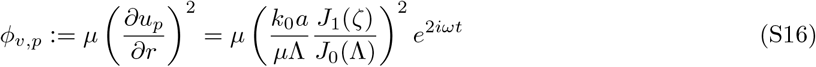

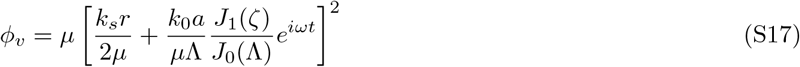

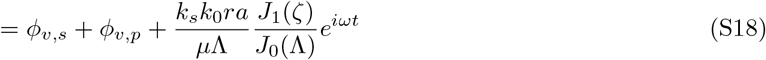

where *ϕ*_*v,s*_ is the VDR contribution from the steady portion of the flow alone, and *ϕ*_*v,p*_ is the VDR contribution from the oscillatory portion of the flow alone, both defined also in Section 2.2. The above simplified expression becomes more complicated once real pulses with multiple frequency contributions are included, where a Fourier decomposition can be used to write down the individual frequency components, and use the expressions above to derive the velocity contributions for each frequency. With these expressions, and with the above derivation, the total VDR *ϕ*_*v*_ can be written as:

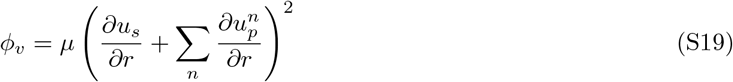

where *n* denotes the number of frequency contributions obtained from Fourier decomposition. The algebraic procedure for obtaining the expressions for each contribution remains the same as demonstrated in the series of equations above.

#### Pulsatile flow in elastic tubes

For the case of pulsatile flow in elastic cylindrical tubes, the generalized expression for the flow velocity, including both the steady and pulsatile contributions is written as:

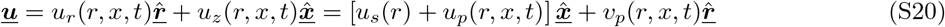

Based on this functional form of the velocity, the contributions from the strain-rate tensor can be written as follows:

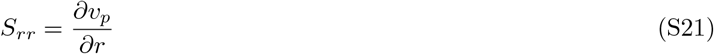

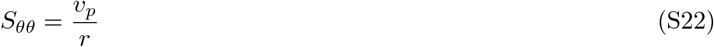

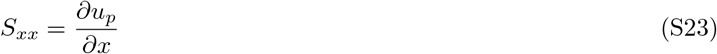

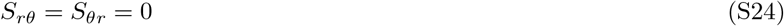

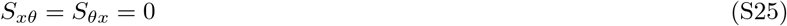

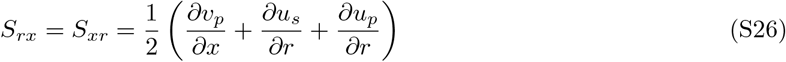

And the resultant total dissipation rate can be obtained as:

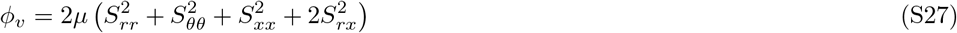

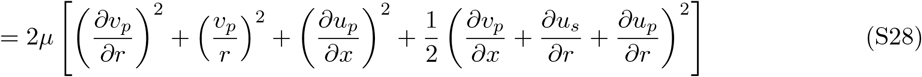

For a purely oscillatory flow contribution, we can set the *u*_*s*_ = 0, which leads to a corresponding VDR contribution from oscillatory flow components alone as follows:

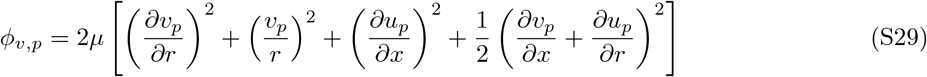

Using the expressions for the steady and oscillatory contribution of velocity as outlined in Section 2.3 now we can obtain a series of final expressions for the various components of the viscous dissipation rate:

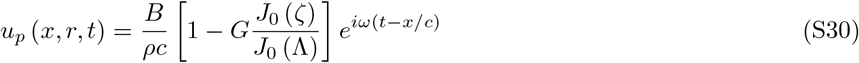

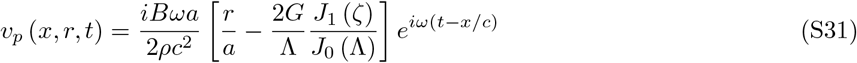

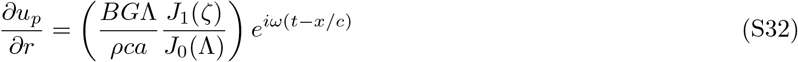

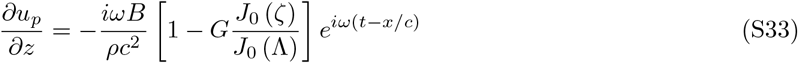

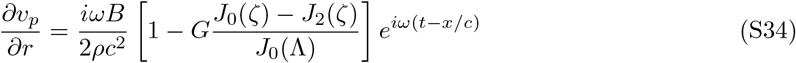

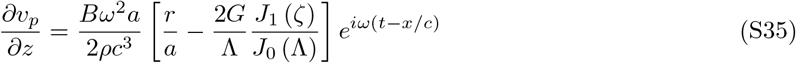

For the case where real pulse waveforms of arbitrary shape are driving the flow, similar to the discussion for rigid tubes, we can decompose the waveform into a steady contribution and a series of oscillatory contributions of varying frequency using a Fourier decomposition. The above expressions can be used to quantify the velocity and the velocity gradients for each frequency contribution, and the resulting total VDR can be written as:

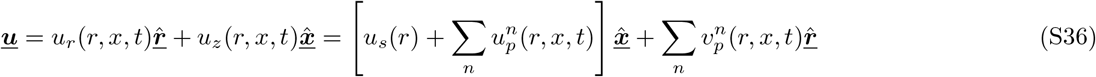

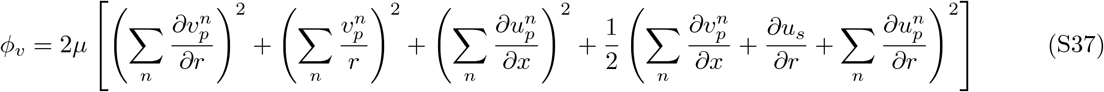

where 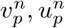 and their derivatines can be quantified using the expressions above; and *n* denotes the summation index across all frequency contributions obtained from the Fourier decomposition. These expressions are sufficiently involved, and hence additional algebraic simplifications are not pursued here. However, with the above forms of the expressions, we can implement them in a calculation scheme using common programming languages such as Matlab and Python.

